# Role of Serum Soluble Interleukin-2 Receptor Level in the Diagnosis of Sarcoidosis: A Systematic Review and Meta-Analysis

**DOI:** 10.1101/2022.07.16.22277713

**Authors:** Samiksha Gupta, Miloni Parmar, Rana Prathap Padappayil, Agam Bansal, Salim Daouk

## Abstract

**Introduction:** Serum Soluble Interleukin-2 Receptor (sIL-2R) levels are used clinically as a disease activity marker for systemic sarcoidosis. Studies have investigated the diagnostic role of serum soluble interleukin-2 receptor (sIL-2R) level for sarcoidosis relative to biopsy. We performed a systematic review and meta-analysis of studies evaluating the diagnostic utility of sIL-2R.

**Methods:** We carried out an electronic search in Medline, Embase, Google Scholar, and Cochrane databases using keyword and Medical Subject Heading (MeSH) terms: sarcoidosis and sIL-2R. Studies evaluating the sIL-2R levels as a diagnostic tool in clinically diagnosed or biopsy-proven sarcoidosis patients compared to control groups with non-sarcoidosis patients were included. Forest plots were constructed using a random effect model depicting pooled sensitivity, specificity, positive and negative predictive values, and diagnostic accuracy.

**Results:** We selected ten studies comprising 1477 patients, with 592 in the sarcoidosis group and 885 in the non-sarcoidosis group. Pooled sensitivity and specificity of sIL-2R levels were 0.88 (95% CI: 0.75-0.95) and 0.87 (95% CI 0.73-0.94) respectively. Pooled negative predictive value and positive predictive value were 0.91 (95% CI 0.77-0.97) and 0.85 (95% CI 0.59-0.96) respectively with diagnostic accuracy of 0.86 (95% CI 0.71-0.93).

**Conclusion:** In addition to its utility as a marker of sarcoidosis disease activity, sIL-2R has high diagnostic accuracy. Despite the limitations of the heterogenous sarcoidosis population and different sIL-2R cutoffs, our results suggest that sIL-2R is an important biomarker that can be used to confirm sarcoidosis diagnosis in unconfirmed or unclear cases.

## Introduction

Sarcoidosis is a heterogenous multi-systemic immune-mediated disease characterized by non-caseating granulomas often localized in the lung or the mediastinal lymph nodes. It is notoriously difficult to diagnose as it is a diagnosis of exclusion and requires an in-depth evaluation of the patient. The diagnosis of sarcoidosis is based on three major criteria: a compatible clinical or radiological presentation, the histological evidence of non-necrotizing granulomatous inflammation in one or more tissues, and the exclusion of alternative causes of this granulomatous disease(1). The absence of a specific diagnostic marker also makes it a challenging diagnosis.

Angiotensin-Converting Enzyme (ACE) is a peptidase secreted by activated macrophages and epithelioid cells within sarcoid granulomas. It is a frequently used laboratory marker for the diagnosis of sarcoidosis, but it lacks sensitivity (22-86%), making it less than helpful in a clinical setting(2,3). In sarcoidosis, the Th1 cytokine pattern seems to predominate mainly in the areas of granuloma formation. Upon activation, Th1 cells upregulate the expression of IL-2R on the cell surface and can shed sIL-2R into circulation(4). sIL-2R levels are therefore used clinically as a disease activity marker for systemic sarcoidosis, as they are an indirect measurement of granuloma burden in the patient(3).

Recently there have been studies evaluating the role of sIL-2R in establishing the diagnosis of sarcoidosis in patients with clinically suspected sarcoidosis. In our review, we aimed to perform a systematic review and meta--analysis of prior studies that evaluated the diagnostic utility of sIL-2R in sarcoidosis.

## Methods

### Literature search

We carried out an extensive electronic search in Medline (PubMed), Embase, Google Scholar, and Cochrane database of systematic reviews, Cochrane central register of controlled trials, Scopus, and Web of science using the keywords/Medical Subject Heading (MeSH) “Sarcoidosis”, “Sarcoid”, “interleukin2 OR interleukin*2*”, “sIL 2r OR sIL-2r”, “il 2 OR IL-2” terms until April 2021 (Supplementary 1 and Supplementary 2). The search also included unpublished articles as well as conference abstracts. The search included articles in all languages.

### Selection of Studies

The studies found on extensive data research were compiled in Covidence software. Covidence software was used for primary and secondary screening by two reviewers. We applied the guidelines of the Preferred Reporting Items for Systematic Reviews and Meta-Analyses (PRISMA) statement to the methods of this study. After removing the duplicated studies, the title and abstracts were independently screened by two authors (SG and RPP). Studies evaluating the sIL-2 receptor levels as a diagnostic tool in clinically diagnosed or biopsy-proven sarcoidosis patients as subjects and the control group with non-sarcoidosis patients were included. Studies done in pulmonary and extra-pulmonary sarcoidosis patients were included. Studies without a control group or addressing only the prognostic performance of the sIL-2 receptor levels were excluded. Studies with less than five patients, including case reports and case series, were excluded. Secondary screening of the included articles was done by two independent authors reviewing the full text (SG and RPP), and the data were extracted. Any discrepancies were resolved by the consensus of the authors.

Further, the reference lists from the retrieved articles were also checked to avoid missing any important studies. Quality Assessment of Diagnostic Accuracy Studies (QUADAS) tool was used to assess the quality of included studies. Two independent authors did the quality assessment.

### Data Extraction

The data was extracted to a Microsoft Excel sheet which included the following variables: type of study, number of assessed patients, number of patients included in sarcoidosis group and control group with non-sarcoidosis patients, age, gender, number of patients with pulmonary and extra pulmonary sarcoidosis, number of patients with a positive sIL-2R in sarcoidosis and non-sarcoidosis group, sensitivity(%), specificity (%), positive and negative predictive value(%).

### Statistical analysis

Forest plots using a random effect model depicting pooled sensitivity, specificity, positive predictive value, negative predictive value, and diagnostic accuracy were constructed. The summary ROC curve was drawn with the calculated area under the curve. Heterogeneity was assessed and reported in I2 and τ2. Data were analyzed using R V.4.0.3.

## Results

After a literature search, there were a total of 796 studies imported from all sources. After removing the duplicate studies, 396 studies were reviewed for abstract screening. Further, 365 studies were excluded on primary screening based on the inclusion and exclusion criteria defined above, and 31 were reviewed for secondary screening. (Figure1). We selected ten studies that fulfilled the inclusion criteria, 6 of which were done in patients with uveitis (Table 1). The cumulative sample size was 1477, with 592 in the sarcoidosis group and 885 in the non-sarcoidosis group. Pooled sensitivity and specificity of sIL-2 receptor levels were 0.88 (95% CI 0.75-0.95) and 0.87 (95% CI 0.73-0.94) respectively. Pooled negative predictive value and positive predictive value were 0.91 (95% CI 0.77-0.97) and 0.85 (95% CI 0.59-0.96) respectively with diagnostic accuracy of 0.86 (95% CI 0.71-0.93) (Table 2). The area under the receiver operating characteristic summary curve was 0.78. On subgroup analysis of patients with uveitis, sensitivity and specificity were 0.84 [0.74-0.96] and 0.79 [0.67-0.93], respectively.

**Table 1:**
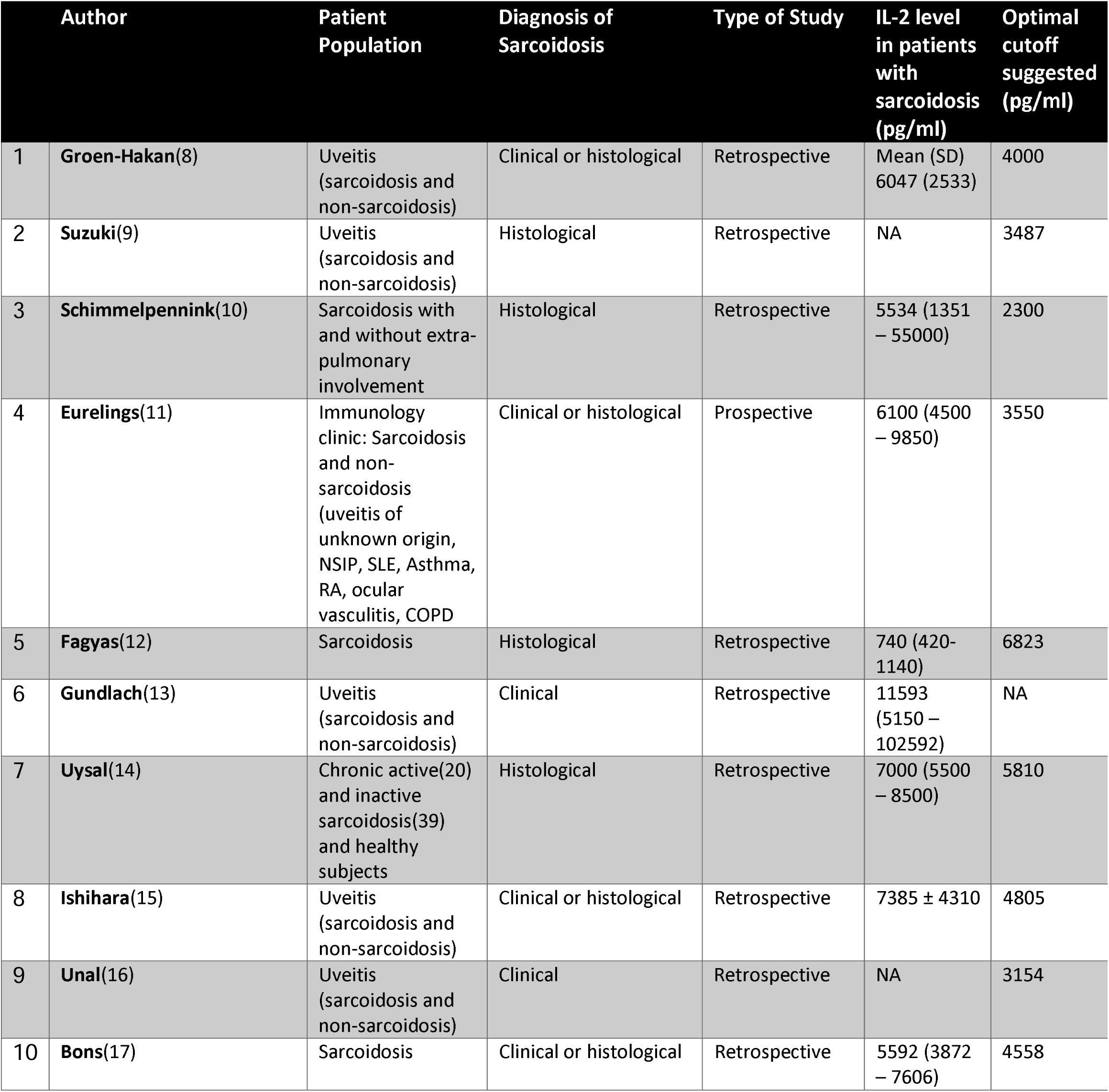
Summary of the included studies

**Table 2:** Summary of the included studies, with first author name, year when article was published (Year), total number of included study subjects(Total), subjects with sarcoidosis(Ns), Subjects without sarcoidosis(Nns), true positives (TP), True negatives (TN), false positives (FP), false negatives(FN), calculated sensitivity and specificity, positive predictive values (PPV), and negative predictive values (PPV)

| S.No | Author | Year | Total | Ns | Nns | TP | TN | FP | FN | Sensitivity | Specificity | PPV | NPV |
| --- | --- | --- | --- | --- | --- | --- | --- | --- | --- | --- | --- | --- | --- |
| 1 | Groen-Hakan | 2017 | 249 | 37 | 212 | 37 | 212 | 119 | 9 | 0.81 | 0.64 | 0.24 | 0.96 |
| 2 | Suzuki | 2020 | 170 | 79 | 91 | 79 | 91 | 15 | 15 | 0.84 | 0.86 | 0.84 | 0.86 |
| 3 | Schimmelpennink | 2020 | 174 | 104 | 70 | 104 | 70 | 0 | 5 | 0.95 | 1 | 1 | 0.93 |
| 4 | Eurelings | 2019 | 189 | 101 | 88 | 101 | 88 | 16 | 14 | 0.88 | 0.85 | 0.86 | 0.86 |
| 5 | Fagyas | 2019 | 104 | 69 | 35 | 69 | 35 | 13 | 64 | 0.52 | 0.73 | 0.84 | 0.35 |
| 6 | Gundlach | 2016 | 247 | 42 | 205 | 42 | 205 | 13 | 1 | 0.98 | 0.94 | 0.76 | 1 |
| 7 | Uysal | 2018 | 84 | 59 | 25 | 59 | 25 | 0 | 0 | 1 | 1 | 1 | 1 |
| 8 | Ishihara | 2020 | 126 | 52 | 74 | 52 | 74 | 6 | 23 | 0.69 | 0.93 | 0.9 | 0.76 |
| 9 | Unal (Letter to the editor) | 2018 | 64 | 14 | 50 | 14 | 50 | 33 | 1 | 0.92 | 0.6 | 0.3 | 0.98 |
| 10 | Bons | 2007 | 70 | 35 | 35 | 35 | 35 | 26 | 21 | 0.62 | 0.57 | 0.57 | 0.63 |

**Figure 1 :**
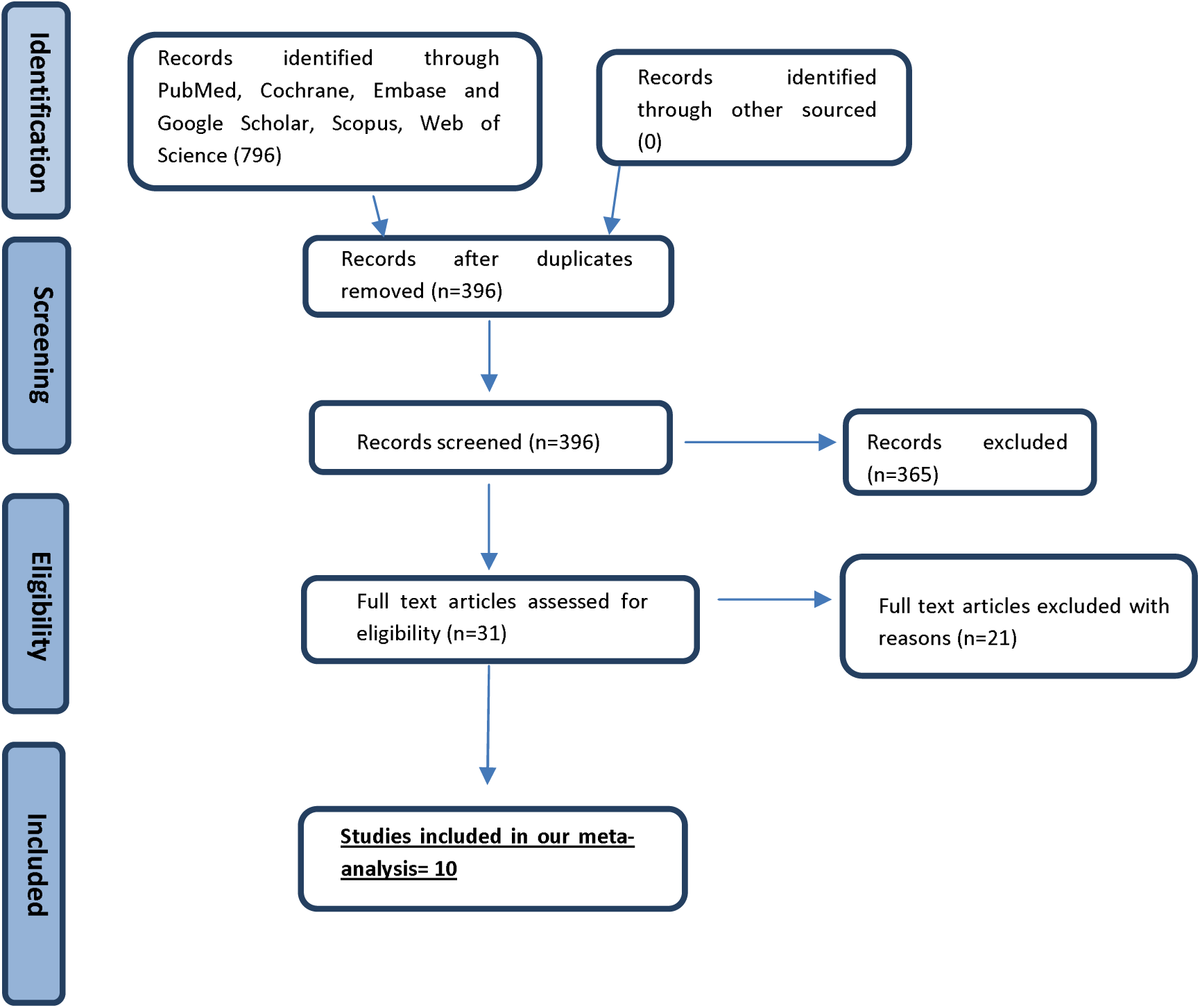
PRISMA flow-chart of inclusion and exclusion of studies in the review.

**Figure 2a :**
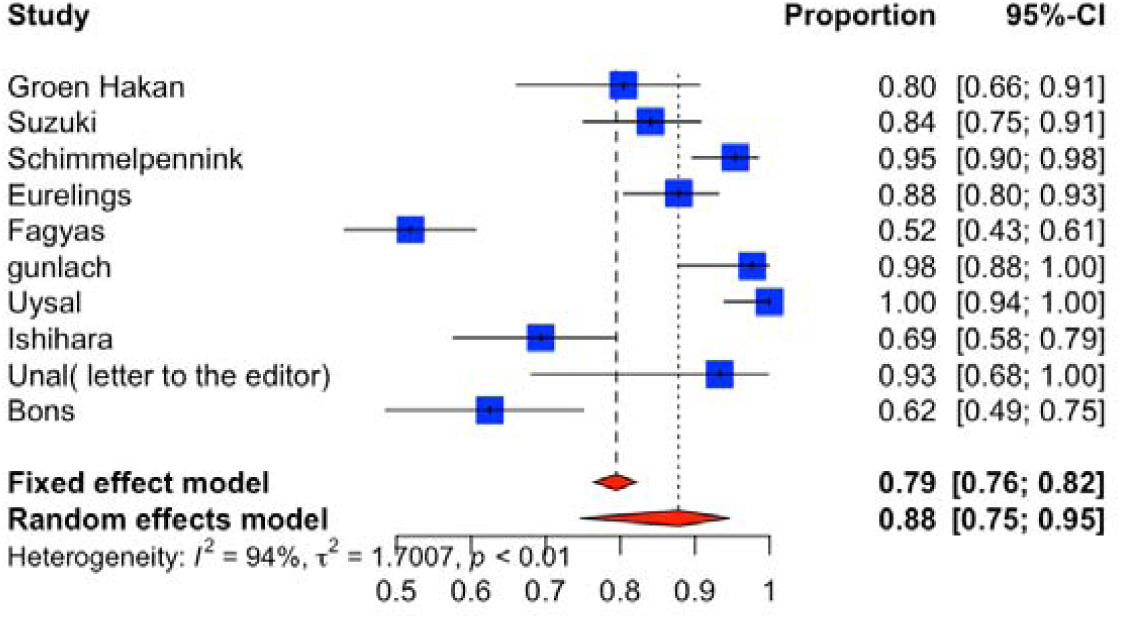
Sensitivity

**Figure 2b :**
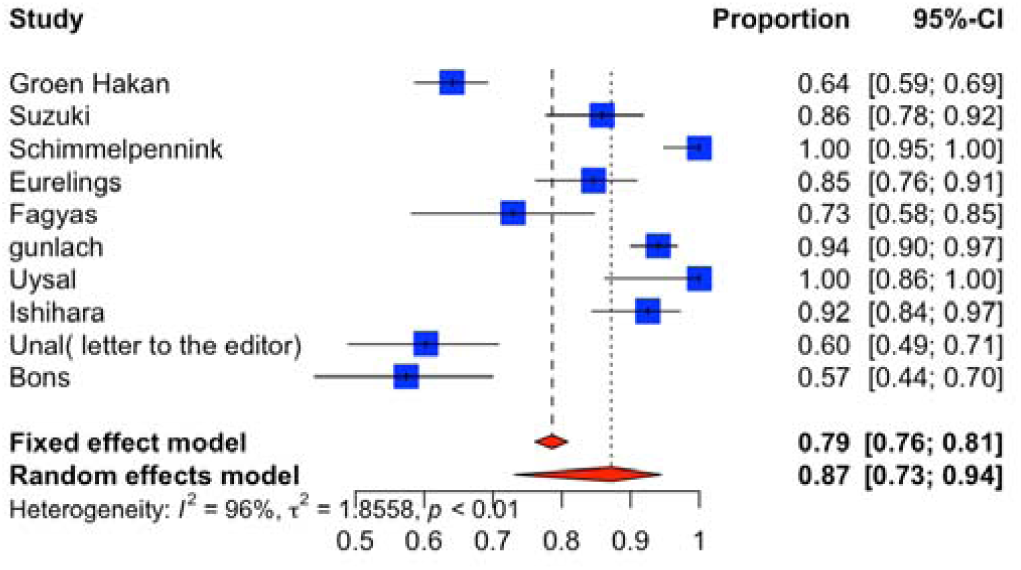
Specificity

**Figure 2c :**
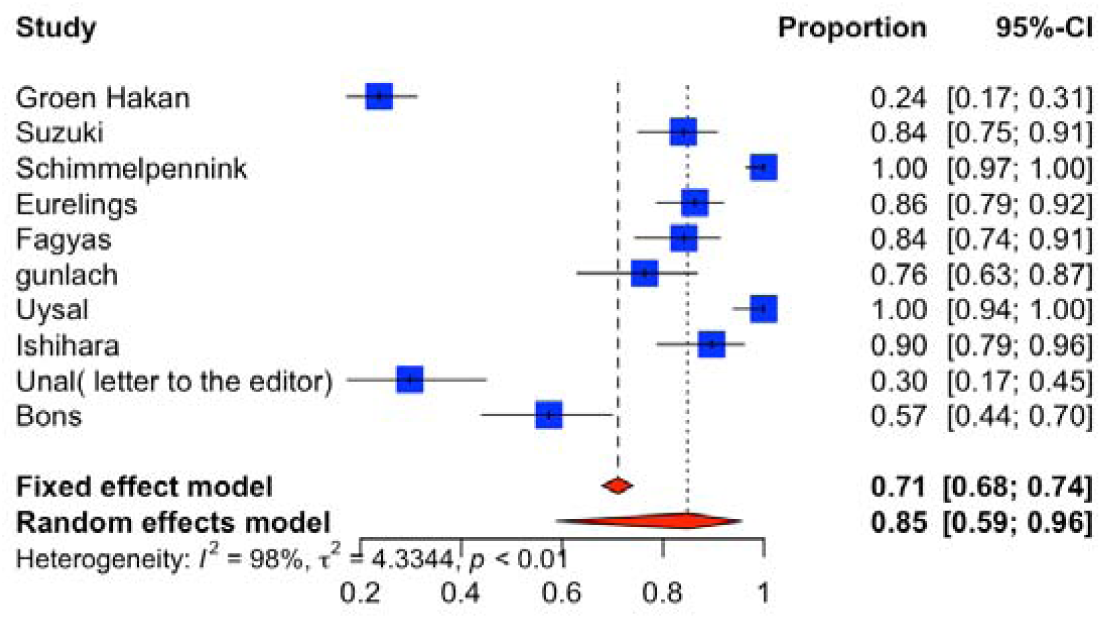
Positive Predictive Value

**Figure 2d :**
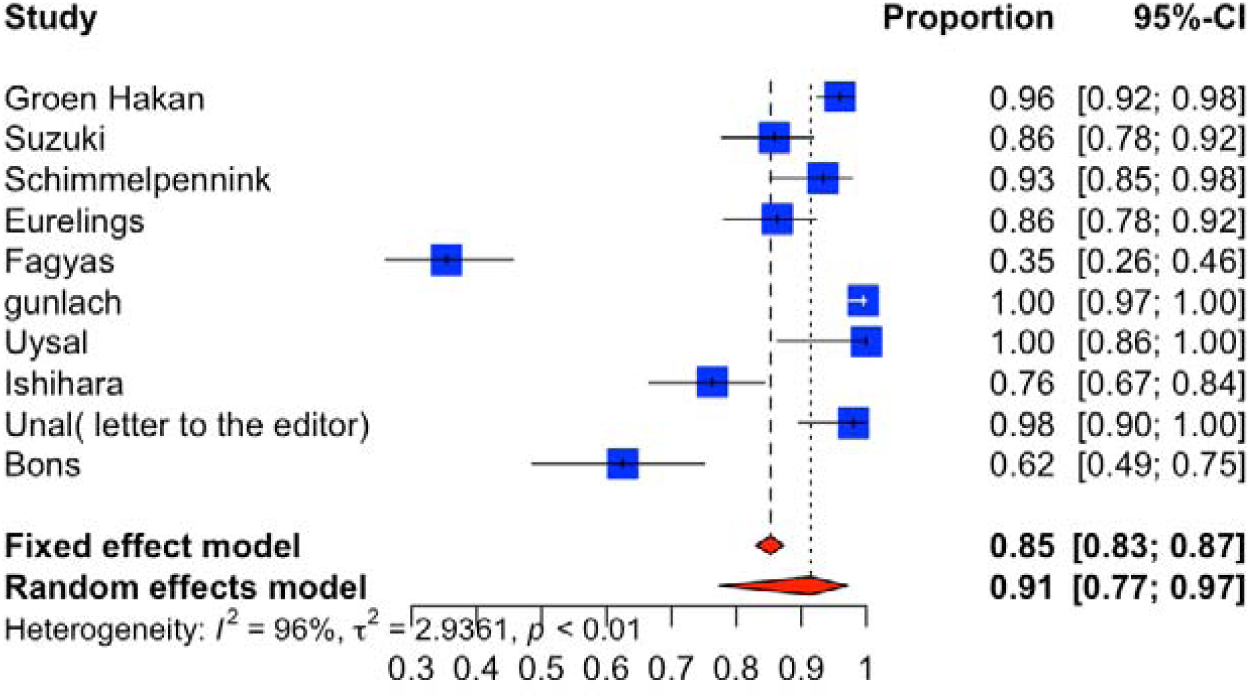
Negative Predictive Value

**Figure 2e :**
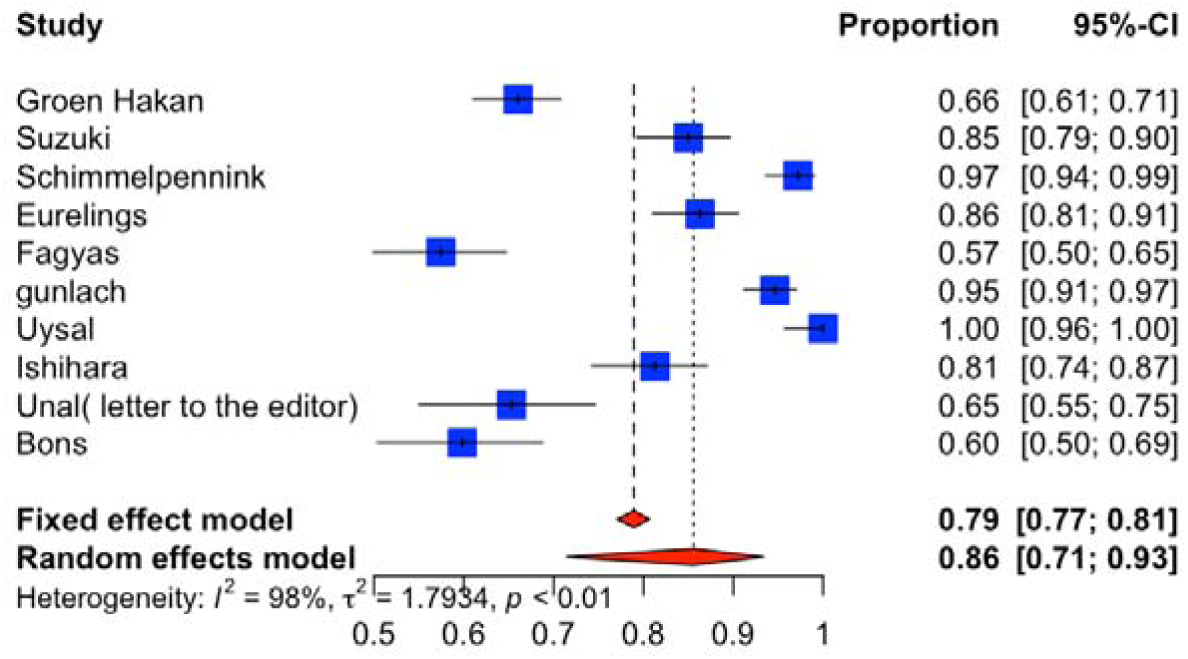
Diagnostic Accuracy

**Figure 2f :**
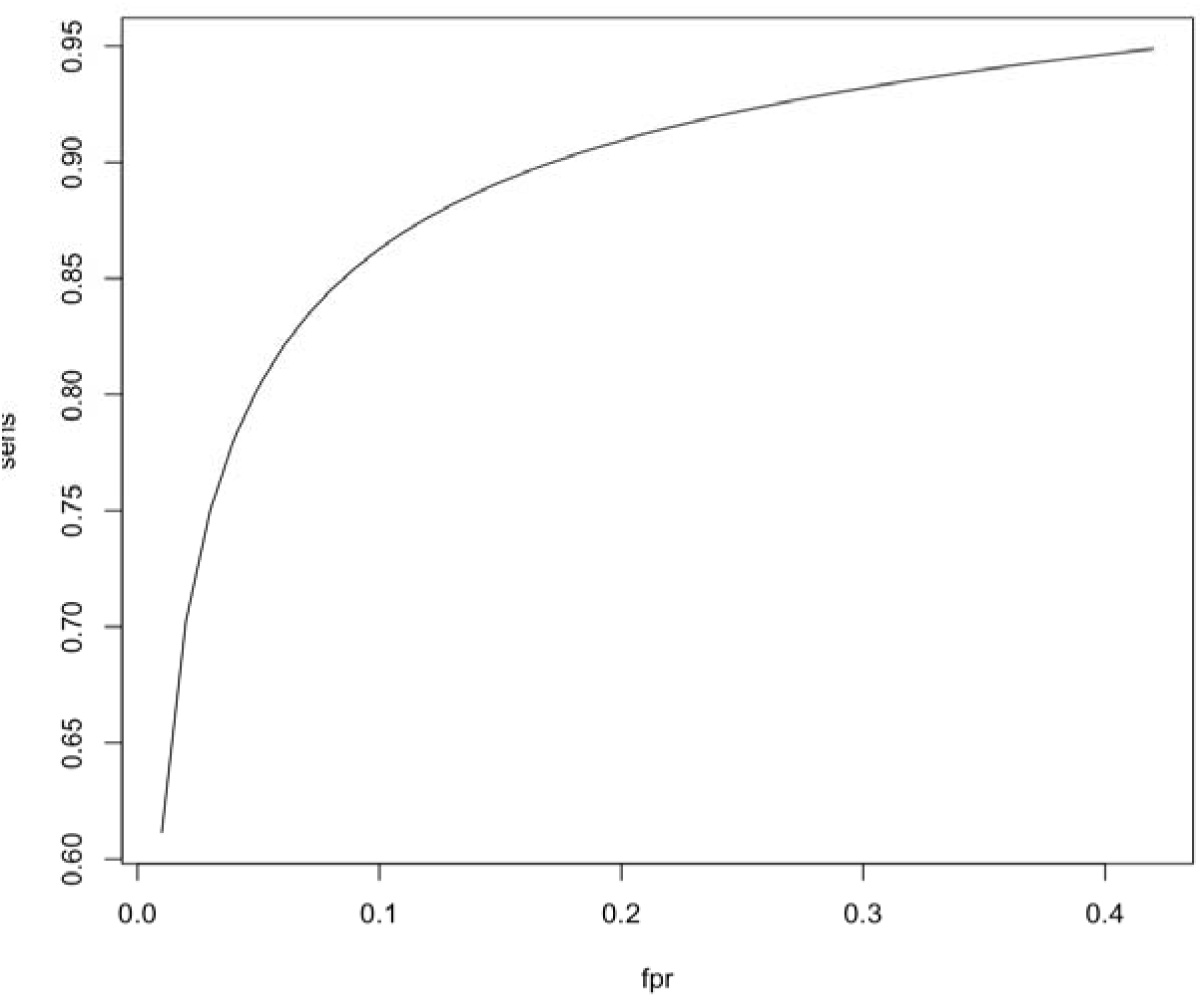
Moses-Shapiro-Littenberg SROC curve (Intercept 3.10, Slope 0.57)

## Discussion

Our systematic review and meta-analysis aimed to assess whether the serum levels of sIL-2 molecules could be used as a potential marker for diagnosing sarcoidosis in patients with suspected clinical sarcoidosis. The diagnostic performance of sIL-2 was also compared by pooling the sensitivity, specificity, positive predictive value, and negative predictive values.

A positive correlation between sIL-2R levels with disease activity in sarcoidosis was demonstrated by Oswald-Richter et al in 2013(5). On similar grounds, Gungor et al. observed that elevated sIL-2R levels were associated with extra-pulmonary involvement(6). Since then, multiple small-scale studies have looked to demonstrate the diagnostic performance of this biomarker. However, these studies primarily suffered from small sample sizes. However, sIL-2R levels showed potential as a biomarker with good diagnostic performance in these studies. Our cumulative review included ten studies and 1477 patients.

Our analysis indicates that sIL-2R levels have a pooled sensitivity of 88% and a specificity of 87%. These numbers are superior to the performance characteristics of previously proposed biomarkers such as ACE levels which have demonstrated sensitivity and specificity of 62% and 76%, respectively (11). The area under the ROC curve is 0.78, which is acceptable for its use as a diagnostic test in the clinical setting. In patients with clinically suspicious sarcoidosis, an elevated sIL-2R level has a high positive predictive value of up to 91%. Based on these findings, we recommend sIL-2R levels be considered an adjuvant in diagnosing patients with clinically suspected sarcoidosis.

The primary limitation of our study is the heterogeneity in the patient population and control groups included in the analyses. The sample sizes of the included studies varied from 64-249, and the study population ranged from patients with ocular sarcoidosis to patients with chronic inactive sarcoidosis. This heterogeneity was also demonstrated in the cutoff of sIL-2R levels used in the studies, which varied from 2300-5800 pg/ml. Since there is no diagnostic gold standard for sarcoidosis, what was considered positive cases in these studies also varied greatly, with not all patients having biopsy-proven sarcoidosis. Hence, even though our observations show great promise for using sIL-2R in a clinical setting, the diagnostic performance of this marker will need to be reassessed in a large-scale study.

Additionally, the clinical adaptation of this test may also come at a financial cost where sIL-2R level may not be readily available or is more expensive in some settings. Levels of sIL-2R can also be affected by factors such as renal function; hence, diagnostic interpretation of these results in a clinical setting should also be cautiously approached (7). Additional factors that would have affected the levels include the variations in the laboratory methods used to assess the sIL-2R levels.

## Conclusion

Our meta-analysis showed that in addition to its utility as a marker of sarcoidosis disease activity, sIL2R has high diagnostic accuracy. Despite the limitations of the heterogenous sarcoidosis population and different sIL2R cutoffs, our results suggest that sIL2R can be used as a more sensitive biomarker compared to the traditional use of ACE, especially to rule out sarcoidosis in unconfirmed or unclear cases.

## Supporting information

Supplemental search statergy

## Data Availability

All data produced in the present work are contained in the manuscript

