## Supplemental search statergy for "Role of Serum Soluble Interleukin-2 Receptor Level in the Diagnosis of Sarcoidosis: A Systematic Review and Meta-Analysis"

**Searches Executed:** May 1, 2021

**Search Sets Forwarded for Review:** Highlighted in each strategy

**Database:** Ovid MEDLINE(R) and Epub Ahead of Print, In-Process, In-Data-Review & Other Non-Indexed Citations and Daily <1946 to April 30, 2021>

**Search Strategy:**

--------------------------------------------------------------------------------

1	exp Sarcoidosis/ (25500)

2	sarcoid$.mp. (32554)

3	1 or 2 (32576)

4	((interleukin$ 2$ or interleukin2$ or il 2 or il2) adj2 receptor$ adj2 solub$).mp. (2945)

5	3 and 4 (125)

6	remove duplicates from 5 (125)

***************************

**Database:** Embase Classic+Embase <1947 to 2021 April 30>

**Search Strategy:**

--------------------------------------------------------------------------------

1	exp sarcoidosis/ (51555)

2	sarcoid$.mp. (55400)

3	1 or 2 (55728)

4	soluble interleukin 2 receptor/ (1496)

5	((interleukin$ 2$ or interleukin2$ or il 2 or il2) adj2 receptor$ adj2 solub$).mp. (4627)

6	4 or 5 (4627)

7	3 and 6 (281)

8	remove duplicates from 7 (277)

***************************

**Database:** EBM Reviews - Cochrane Central Register of Controlled Trials <March 2021>

**Search Strategy:**

--------------------------------------------------------------------------------

1	exp sarcoidosis/ (216)

2	sarcoid$.mp. (738)

3	1 or 2 (738)

4	soluble interleukin 2 receptor/ (0)

5	((interleukin$ 2$ or interleukin2$ or il 2 or il2) adj2 receptor$ adj2 solub$).mp. (265)

6	4 or 5 (265)

7	3 and 6 (3)

8	remove duplicates from 7 (3)

***************************

**Database:** EBM Reviews - Cochrane Database of Systematic Reviews <2005 to April 28, 2021>

**Search Strategy:**

--------------------------------------------------------------------------------

1	sarcoid$.af. (74)

2	((interleukin$ 2$ or interleukin2$ or il 2 or il2) adj2 receptor$ adj2 solub$).af. (22)

3	1 and 2 (0)

***************************
